# Safety, tolerability and immunogenicity of Biological E’s CORBEVAX™ vaccine in children and adolescents: A Prospective, Randomised, Double-blind, Placebo controlled, Phase-2/3 Study

**DOI:** 10.1101/2022.04.20.22274076

**Authors:** Subhash Thuluva, Vikram Paradkar, SubbaReddy Gunneri, Vijay Yerroju, Rammohan Mogulla, Pothakamuri Venkata Suneetha, Kishore Turaga, Mahesh Kyasani, Senthil Kumar Manoharan, Srikanth Adabala, Aditya Sri Javvadi, Guruprasad Medigeshi, Janmejay Singh, Heena Shaman, Akshay Binayke, Aymaan Zaheer, Amit Awasthi, Manish Narang, Pradeep Nanjappa, Niranjana Mahantshetti, Bishan Swarup Garg, Mandal Ravindra Nath Ravi

## Abstract

**Background:** After establishing safety and immunogenicity of Biological E’s CORBEVAX™ vaccine in adult population (18-80 years) in Phase 1-3 studies, vaccine is further tested in children and adolescents in this study.

**Methods:** This is a phase-2/3 prospective, randomised, double-blind, placebo controlled, study evaluating safety, reactogenicity, tolerability and immunogenicity of CORBEVAX™ vaccine in children and adolescents of either gender between <18 to ≥12 years of age in Phase-II and <18 to ≥5 years of age in Phase-III with placebo as a control. This study has two age sub groups; age subgroup-1 with subjects <18 to ≥12 years of age and age subgroup-2 with subjects <12 to ≥5 years of age. In both age sub groups eligible subjects (SARS-CoV-2 RT-PCR negative and seronegative at baseline) were randomized to receive either CORBEVAX™ vaccine or Placebo in 3: 1 ratio.

**Findings:** The safety profile of CORBEVAX™ vaccine in both pediatric cohorts was comparable to the placebo control group. Majority of reported adverse events (AEs) were mild in nature. No severe or serious AEs, medically attended AEs (MAAEs) or AEs of special interest (AESI) were reported during the study period and all the reported AEs resolved without any sequelae. In both pediatric age groups, CORBEVAX™ vaccinated subjects showed significant improvement in humoral immune-responses in terms of anti-RBD-IgG concentrations, anti-RBD-IgG1 titers, neutralizing antibody (nAb)-titers against Ancestral Wuhan and Delta strains. Significantly high interferon gamma immune response (cellular) was elicited by CORBEVAX™ vaccinated subjects with minimal effect on IL-4 cytokine secretion.

**Interpretations:** The safety profile of CORBEVAX™ vaccine in <18 to ≥5 years’ children and adolescents was found to be safe and tolerable. The adverse event profile was also found to be acceptable. Significant increase in anti-RBD IgG and nAb titers and IFN-gamma immune responses were observed post vaccination in both pediatric age sub groups. Both humoral and cellular immune responses were found to be non-inferior to the immune responses induced by CORBEVAX™ vaccine in adult population. This study shows that CORBEVAX™ vaccine is highly immunogenic and can be safely administered to pediatric population as young as 5 years old.

The study was prospectively registered with clinical trial registry of India-CTRI/2021/10/037066

## INTRODUCTION

COVID-19 infection among children is generally asymptomatic or mild in nature compared to adults. However, severe COVID-19 associated cases of multi system inflammatory syndrome (MIS-C) and long-term sequelae, such as the long COVID, cardiovascular complications, neurological manifestations, dermatological manifestations, acute kidney failure etc. were reported in children.^1-4^ Importantly, SARS-CoV-2 can be efficiently transmitted from school-age children and adolescents to household members and can led to the hospitalization of adults with secondary cases of Covid-19.^5^ Moreover, home confinement, isolation during the infection and reduced physical activity had greater impact on their lifestyle, psychological and social well-being.^6,7^ Infection transmission chain from childrens’ silent infections cannot be contained solely based on vaccination of adult population. So, there is an urgent medical need of immunizing the children with COVID-19 vaccine. Though many vaccines tested on different platforms are approved in adult population, very few clinical studies were conducted in pediatric population. BNT162b2 Pfizer-BioNTech is available globally for vaccinating the 5-17 years aged population. Some countries have given emergency use authorization (EUA) to mRNA vaccines (Pfizer and Moderna) for use in the adolescent group (12-17 years). Both Pfizer BioNTech and Moderna vaccines are reported to be well tolerated and shown 99-100% efficacy in population aged between 12-25 years^8^ and 12-17 years^9^ respectively. Two inactivated vaccines (Sinovac-CoronaVac and BBIBP-CorV) were approved by Chinese authorities for the age group of 3-17 years.^10,11^ An adjuvanted inactivated vaccine (Covaxin) and a novel DNA vaccine, ZycovD were approved by the Indian regulators in the 12-17 years age group, but not yet received WHO EUL for this age group. Till date, there is only one phase-1/2 clinical trial data available testing safety of the protein sub-unit vaccine in children^12^ and no studies were published on establishing the safety and immunogenicity of protein sub unit COVID-19 vaccines from large pivotal phase-3 trials.

CORBEVAX™ is a protein sub unit vaccine containing receptor binding domain (RBD) of spike protein of SARS-CoV-2, adjuvanted with CpG1018 and Al3+. Safety and immunogenicity of CORBEVAX™ is well established in our previous Phase 1-3 clinical trials in adult population.^13^ In Adults, vaccine exhibited excellent safety and minimal reactogenicity with no observed safety concerns. Neutralizing antibody titers in CORBEVAX™ vaccinated subjects indicated high vaccine effectiveness in preventing symptomatic infection and severe disease; >90% vaccine effectiveness against Wuhan variant and >80% vaccine effectiveness against the Delta variant based on neutralizing antibodies correlates of protection.^13^ In the study, we further report safety and immunogenicity of CORBEVAX™ in a prospective, randomised, double-blind, placebo controlled, phase-2/3 study conducted in children and adolescents.

## METHODS

### Study Design and Study Population

This is an ongoing prospective, randomized, double-blind, placebo controlled, seamless phase-2/3 study to demonstrate safety, reactogenicity & immunogenicity of BE’s CORBEVAX™ vaccine in children and adolescents. Study was conducted at 23 sites across India in accordance with the principles defined in the Declaration of Helsinki, International Conference on Harmonization guidelines (Good Clinical Practices), and the local regulatory guidelines. The Investigational Review Board or Ethics Committee at each study site approved the protocol. All participants provided written informed consent before enrollment into the study. Complete list of eligibility criteria provided as supplementary information.

Participants were healthy children (SARS-CoV-2 RT-PCR and anti-SARS-CoV-2 antibody negative) between <18 to ≥5 years of age. The enrolment was done in an age descending approach with two age subgroups viz. age subgroup-1 with subjects <18 to ≥12 years of age and age subgroup-2 with subjects <12 to ≥5 years of age. Study subjects were randomized to receive either test vaccine (CORBEVAX™) or placebo in a 3:1 ratio in both age subgroups. A total of 1910 subjects were screened to enroll 624 (n=312 in age sub groups 1 and n=312 in age sub group 2) subjects into the study. All enrolled subjects received either two doses of CORBEVAX™ vaccine or placebo through intramuscular route on Day0 and Day28(+4). From age sub group-1, 134 subjects were enrolled into phase-2 part of the study (100 participants were randomized to receive CORBEVAX™ vaccine and 34 subjects received placebo). Safety data from phase-2 study participants was reviewed by Data Safety Monitoring Board (DSMB) at day-7 post first dose of vaccination. Based on the favorable recommendations by DSMB on safety profile of the CORBEVAX™ vaccine in adolescents, study was progressed to phase-3 and recruited remaining 178 subjects in age sub group-1 (134 subjects, under test vaccine arm and 44 subjects under placebo arm) and n=312 subjects were recruited in age sub group-2, 5-11 years old children (234 subjects, under test vaccine arm and 78 subjects under placebo arm). During the conduct of this study, there were no major protocol deviations reported at any of the study sites. Few subjects reported for their visits out of window period but these deviations were not found to be significant and all deviations were communicated to ethics committees of the respective study sites.

### Study Duration

The total duration of the study (interim analysis) was 56 days for each subject from the 1^st^ dose of vaccination and are followed up further for 3 months and 6 months after the second dose. A booster dose will be given at 6 months post 2^nd^ dose visit and subjects will be followed up for 28 days’ post booster dose. A time window of +4 days is allowed from Day 28 to Day 56 and a time window of 14 days will be allowed for 3 months and 6 months follow up respectively to ensure participant compliance.

### Randomization and masking

After all screening-related activities were completed and prior to the first dose of study vaccine, participants in the study were stratified and randomized to either test or placebo groups in 3:1 ratio according to a random generated codes using the interactive web response system (IWRS) platform. Participants and investigators were blinded to group allocation. Except independent statistician and personnel responsible for conducting packaging/ labelling/blinding of the investigational product, all the personnel involved in the study remained blinded to the study vaccine. The test and placebo were labelled and packed identical to each other.

### Procedure

Biological E’s CORBEVAX™ vaccine is a recombinant sub unit vaccine consists of RBD protein as an antigen, CpG 1018 and Aluminium hydroxide as adjuvants formulated in Tris buffer.^13^ The recombinant RBD protein is produced in Pichia Pastoris culture as secretory protein and purified via multiple chromatography and ultrafiltration/normal-filtration steps. A 0.5mL of CORBEVAX™ or placebo (Adjuvant formulated in Tris-buffer) was administered via an intramuscular (IM) injection into the deltoid muscle in a 2-dose schedule (Day 0 & Day 28). Prophylactic medication is not prescribed either before or after vaccination.

Participants were evaluated with SARS-CoV-2 real-time RT-PCR and sero-status was assessed using Anti-SARS CoV-2 Human S1/S2 IgG ELISA using Diasorin kits. Participants who were negative for both SARS CoV-2 infection and Anti-SARS CoV-2 human S1/S2 IgG antibodies were enrolled into the study.

### Outcomes

The primary outcome of the phase-2 and secondary outcome of phase-3 part of the study was to assess the safety, tolerability and reactogenicity of CORBEVAX™ vaccine in children and adolescents in comparison to placebo group in terms of occurrence of any solicited and unsolicited AEs for 28 days’ post vaccination. The primary outcome of phase-3 part of the study was demonstration of immunogenic non-inferiority of BE’s CORBEVAX™ vaccine against adult population in terms of geometric mean neutralizing titers and their geometric mean fold rise from baseline, at day 42 (14 days after 2nd dose). Exploratory outcomes include measuring, SARS-COV-2 virus neutralizing antibodies against variant strains (Beta and Delta), anti-RBD IgG subclass (IgG1 and IgG4) titer assessment, cellular immune response assessment at baseline and at Day 42 in a randomly selected subset of study population. Other exploratory outcomes were to assess occurrence and severity of SAEs and medically attended AEs (MAAEs) and persistence of immune responses at 3 and 6 months’ post 2^nd^ dose of vaccination.

### Safety assessments

Occurrence and severity of all adverse reactions were collected for the safety assessments, which includes solicited and unsolicited, non-serious and serious adverse events (AEs), medically attended AEs (MAAEs) and adverse events of special interest (AESI) reported in the study from the time of first dose of the vaccine. All participants were observed for at least 60 mins post vaccination to assess reactogenicity. Solicited local and general AEs were recorded for 7 consecutive days (Day 0-6), captured through subject diary after each vaccine dose. Unsolicited AEs, if any were collected until day 42. SAEs, MAAEs and AESI were collected throughout the study period. Relatedness of study vaccine was assessed for all reported AEs and adverse reactions were scored by severity (mild, moderate, severe and life threatening). Details on assessment of reported AEs was provided as supplementary information.

### Immunogenicity Analysis

Sera samples were collected from all the subjects in the immunogenicity cohort at Day-0 (pre-vaccination) and at Day-42 (14 days after second vaccine dose) time points. Following measurements were conducted to assess humoral immune response

1. Overall humoral immune response generated by vaccination was tested by measuring the anti-RBD IgG concentration via ELISA method conducted at Dang’s Lab, India. Subject sera samples collected prior to vaccination (Day-0) and fourteen days after two doses of vaccination (Day-42). The antibody concentrations were reported in ELISA Units/mL for each subject and Geometric Mean Concentrations were calculated for both age sub group cohorts at day0 and day42.
2. Th1 vs Th2 skew of humoral response was assessed by measurement of anti-RBD IgG1 and IgG4 isotype titers for Day-0 and Day-42 time-point via ELISA method conducted at Dang’s Lab, India.
3. Neutralizing Antibody titers were tested in the subject sera samples collected prior to vaccination (Day-0) and fourteen days after two doses of vaccination (Day-42). The testing was conducted against Wild-Type SARS-COV-2 strains in a Micro Neutralization Assay (MNA) in a BSL-3 facility at THSTI, India. MNA assay originally developed and validated at PHE, UK laboratory and then transferred and revalidated at THSTI. For prototype Wuhan Strain, the Victoria isolate from USA was used while the Delta strain used in the assay was isolated in India.
4. Cellular immune response involved direct measurement of cytokines secreted by stimulation of whole blood samples with SARS-COV-2 peptides. So, whole blood samples collected at the clinical trial site were directly incubated in specialized tubes that are coated with SARS-COV-2 peptides in the presence of growth medium. Concentrations of Interferon-gamma and Interleukin-4 cytokines recorded at Day-0 and Day-42 time-points were then tested by standard BD ELISA kits at Dang’s Lab. The total Interferon-gamma secreting PBMC’s was also assessed by ELISPOT method conducted at THSTI, India. Whole blood samples were collected post two-dose vaccination and PBMC’s were isolated and stored frozen. The PBMC’s were subsequently stimulated with various stimulants; SARS-COV-2 RBD peptides for specific response, DMSO for non-specific response and PHA for assay validity criteria. Post-stimulation, the number of PBMC’s that secrete cytokine Interferon-gamma were identified and quantified by ELISPOT technique and the Spot Forming Units (SFU’s) per million PBMC’s were calculated for each subject sample. Additional information in Supplementary section.

### Statistical Analyses

There are two pediatric subgroups stratified by age viz <18 to ≥12 years, <12 to ≥5 years in this study. Sample size was sufficiently powered at subgroup level to allow for separate comparisons of geometric mean titers at day 42 for each of the two pediatric subgroups against the adult immunogenicity data.

Immunogenic non-inferiority of BE’s CORBEVAX™ vaccine in pediatric population was demonstrated against adult population in terms of geometric mean neutralizing titers and their geometric mean fold rise from baseline, at day 42. Data of adult cohort was used from another phase-2/3 study^13^ (BECT-069) which was conducted prior to the start of the pediatric study and formed the basis for non-inferiority comparison. Non-inferiority (NI) was assessed separately for each subgroup using the confidence interval for the GMT ratio method. For sample size estimation the true GMT ratio was assumed to be 0.8 with a standard deviation of log transformed data to be 1.2. The lower limit of NI margin is set at 0.5. The non-inferiority will be inferred if the lower limit of two sided 95% CI of GMT ratio is ≥0.5 limit set. The sample size of 210 evaluable subjects (SAS 9.4) in each age group will give around 87% power at 5% two-sided level of significance to test the GMT ratio between each age group with adult sample data. Including 10% dropout allocation, the required sample size will then be 234 in each of the two age groups. Accordingly, the overall sample size in this phase-2/3 study would then be 468 eligible children. The overall sample size would be 624 subjects.

Immunogenicity was compared using a two-sample t-test on the means of log-transformed titers at the 5% significance level in order to allow for pairwise comparisons. For each comparison, the ratio of the GMTs along with their corresponding 95% CI was presented.

For the purposes of analysis, recruited subjects were further identified as total vaccinated cohort (TVC) for safety assessment and the according to protocol (ATP) cohort for immunogenicity assessment. All the demographic and primary safety analyses have been based on TVC population, defined as subjects who entered into the study and have received at least one single intramuscular dose of study vaccination or placebo. ATP population is defined as population, who have blood samples available for immunogenicity analysis at all protocol specified time points from CORBEVAX™ vaccinated cohorts.

All data were summarized descriptively and data listings were based on all subjects enrolled in the study. By default, descriptive statistics for quantitative measurements included the number of subjects (n), mean, standard deviation (SD), minimum, median (IQR) and maximum. Safety data were summarized by System Organ Class and Preferred Term. Serious adverse events, related adverse events, adverse events leading to death or withdrawal, solicited adverse events, MAAEs and AESI were summarized separately. In addition, adverse events were also summarized by severity. All analyses were conducted using SAS® Version 9.4 or higher.

### Role of the funding source

BIRAC-a division of the Department of Biotechnology, Govt of India provided partial funding for the execution of trials. CEPI provided support for nAb titer testing in terms of reagents. Funding sources were not involved in the study conduct, data analysis/interpretation or writing the manuscript.

## RESULTS

### Participants

In total, 1910 subjects were screened and 312 eligible subjects were enrolled into <18 to ≥12-year group and another 312 subjects were enrolled into <12 to ≥5-year group. Overall, 11 subjects dropped out from the study, 4 from age sub group-1 and 7 from age sub group-2 due to lost to follow up and migration from study area (Fig 1). All the 624 subjects received two doses (0.5 ml) of either Biological E’s CORBEVAX™ vaccine (468 subjects) or Placebo (156 subjects) and were part of safety analyses. Demographics of total vaccinated group were presented in table 1.

**Figure 1:**
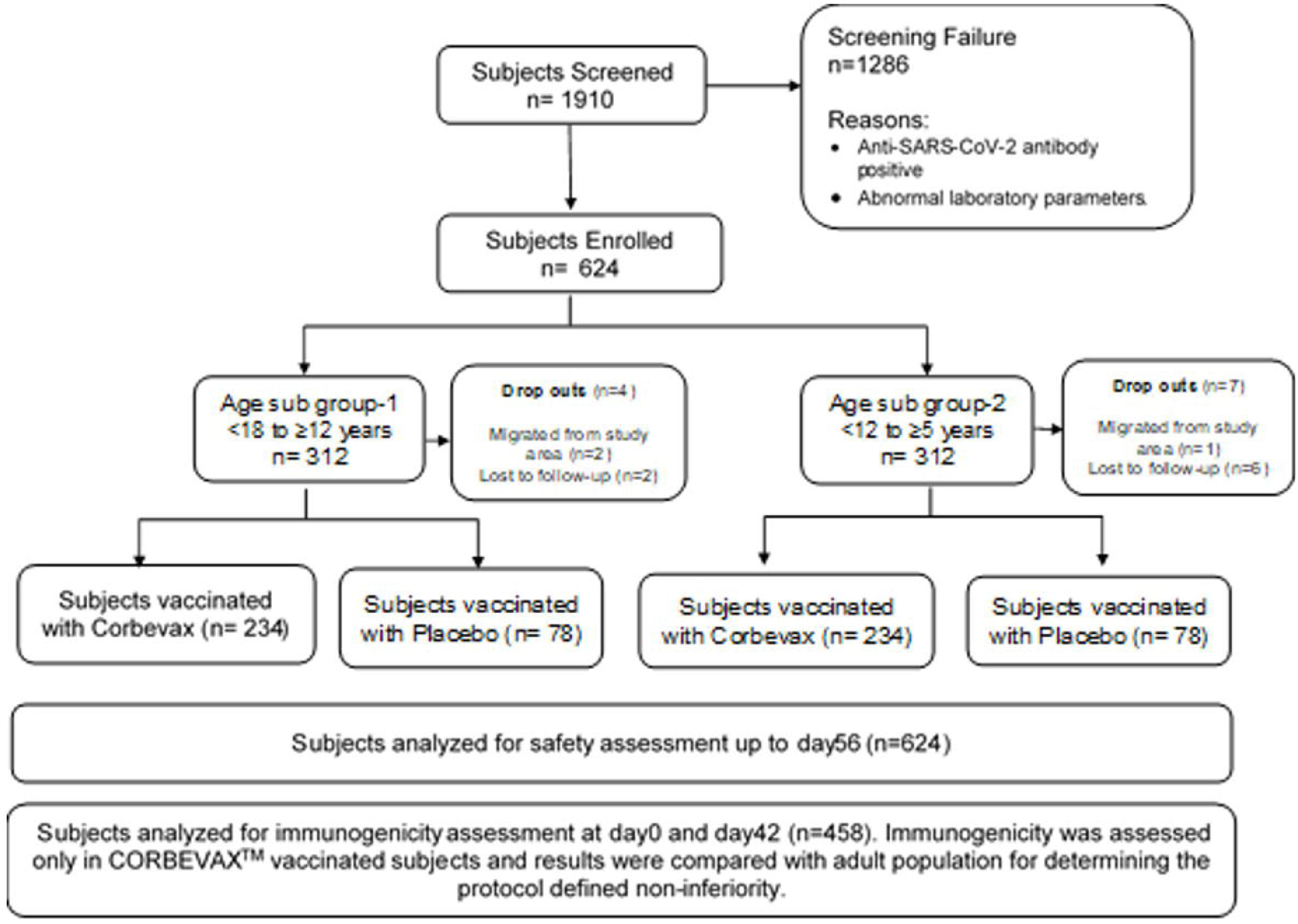
Subject disposition: A total of 1910 subjects were screened and 624 eligible subjects were enrolled into the study. Out of 624 subjects, 312 subjects were enrolled in <18 to ≥ 12 years age sub group and randomized to either CORBEVAX™ vaccination group or placebo group in a 3:1 ratio. Similar approach was followed for <12 to 5 years age sub group cohort (n=312). All subjects were of Indian origin and male, female ratio was balanced in both age sub groups. All the 624 subjects received two doses (0.5 ml) of either Biological E’s CORBEVAX™ vaccine (468 subjects) or placebo (156 subjects) intramuscularly in the deltoid muscle of the non-dominant upper arm and completed their follow up period of Day-56. A total of 11 subjects dropped out from the study, either due to migration from study area or due to lost to follow up. All 624 subjects were assessed for safety analyses and 458 subjects out of 468 (97.8%) subjects were included in the immunogenicity assessment.

**TABLE 1:** Demographic characteristics of study participants.

| Demographic-Variables<br>Statistics / Category | <18 to ≥12 years | <12 to ≥5 years |
| --- | --- | --- |
|  | N=312 | N=312 |
|  | n (%) | n (%) |
| Age (Years) |  |  |
| n | 312 | 312 |
| Mean | 14.7 | 8.8 |
| Nationality |  |  |
| Indian | 312 (100.00) | 312 (100.00) |
| Gender |  |  |
| Male | 162 (51.92) | 169 (54.17) |
| Female | 150 (48.08) | 143 (45.83) |

Percentages were calculated using column header group count as denominator.
n: Subject count in specified category. N= Total number of subject.

### Safety Findings

All 624 enrolled subjects were followed up until day 56 post first dose of vaccination for any solicited local and systemic adverse reactions. 234 subjects in each age sub group were vaccinated with CORBEVAX™ and 78 subjects in each age sub group were vaccinated with placebo.

In <18 to ≥12 years (n=312) sub group, 66 (28.2%) subjects reported 118 events under CORBEVAX™ arm (n=234) and 22 (28.2%) subjects reported 35 events under Placebo arm (n=78). In <12 to ≥5 years (n=312) sub group, 52 (22.2%) subjects reported 95 events under CORBEVAX™ arm (n=234) and 26 (33.3%) subjects reported 45 events under Placebo arm (n=78). Overview of the AEs reported in the study were shown in table 2. Summary of AEs by SOC and PT in TVC group were presented in table 3. All these adverse events were solicited in nature in <18 to ≥12 years sub group. Two unsolicited AEs reported in CORBEVAX™ vaccinated group and one reported in placebo group in <12 to ≥5 years’ age sub group cohort.

**Table 2:** Overview of subjects with AEs – Total Vaccinated Group.

| Category<br>N1 (%) n | <18 to ≥12 years (n=312) |  | <12 to ≥5 years (n=312) |  |
| --- | --- | --- | --- | --- |
|  | CORBEVAX™ | PLACEBO | CORBEVAX™ | PLACEBO |
|  | N=234 | N=78 | N=234 | N=78 |
| Any AE | 66 (28.2) 118 | 22 (28.2) 35 | 52 (22.2) 95 | 26 (33.3) 45 |
| Any Serious AE | 0 (0.0) 0 | 0 (0.0) 0 | 0 (0.0) 0 | 0 (0.0) 0 |
| Any Local AE | 44 (18.8) 57 | 18 (23.1) 26 | 37 (15.8) 49 | 22 (28.2) 32 |
| Any Systemic AE | 36 (15.4) 61 | 8 (10.3) 9 | 29 (12.4) 46 | 10 (12.8) 13 |
| Any Medically Attended AE | 1 (0.4) 2 | 0 (0.0) 0 | 0 (0.0) 0 | 0 (0.0) 0 |
| Any AE within 60 minutes postvac | 14 (6.0) 26 | 2 (2.6) 2 | 8 (3.4) 16 | 6 (7.7) 8 |
| Any Solicited AE | 66 (28.2) 118 | 22 (28.2) 35 | 51 (21.8) 92 | 26 (33.3) 44 |
| Any Unsolicited AE | 0 (0.0) 0 | 0 (0.0) 0 | 3 (1.28) 3 | 1 (0.0) 1 |

Percentages were calculated using column header count as denominator.
N<sub>1</sub>: Subject Count, N: Sample Size, n:Event Count

**Table 3:** Summary of AEs by SOC and PT in subjects vaccinated with CORBEVAX™ vaccine.

| SOC/PT<br>N<sub>1</sub> (%) n | <18 to ≥12 years (n=312) |  | <12 to ≥5 years (n=312) |  |
| --- | --- | --- | --- | --- |
|  | CORBEVAX™ | PLACEBO | CORBEVAX™ | PLACEBO |
|  | N=234 | N=78 | N=234 | N=78 |
| OVERALL | 66 (28.2) 118 | 22 (28.2) 35 | 52 (22.2) 95 | 26 (33.3) 45 |
| Gastrointestinal disorders | 3 (1.3) 4 | 0 (0.0) 0 | 2 (0.9) 2 | 2 (2.6) 2 |
| Abdominal pain | 0 (0.0) 0 | 0 (0.0) 0 | 1 (0.4) 1 | 0 (0.0) 0 |
| Nausea | 3 (1.3) 4 | 0 (0.0) 0 | 1 (0.4) 1 | 1 (1.3) 1 |
| Tooth Ache | 0 (0.0) 0 | 0 (0.0) 0 | 0 (0.0) 0 | 1 (1.3) 1 |
| General disorders and administration site conditions | 61 (23.6) 94 | 21 (26.9) 31 | 51 (21.8) 81 | 26 (33.3) 40 |
| Chills | 5 (2.1) 6 | 0 (0.0) 0 | 2 (1.5) 2 | 0 (0.0) 0 |
| Fatigue | 1 (0.4) 1 | 0 (0.0) 0 | 3 (1.3) 3 | 0 (0.0) 0 |
| Injection site erythema | 6 (2.6) 6 | 3 (3.8) 4 | 8 (3.4) 11 | 6 (7.7) 7 |
| Injection site pain | 38 (16.2) 45 | 17 (21.8) 21 | 30 (12.8) 34 | 17 (21.8) 20 |
| Injection site swelling | 6 (2.6) 6 | 1 (1.3) 1 | 4 (1.7) 4 | 4 (5.1) 5 |
| Pyears'exia | 27 (11.5) 30 | 5 (6.4) 5 | 23 (9.8) 27 | 7 (9.0) 8 |
| Musculoskeletal and connective tissue disorders | 8 (3.4) 9 | 0 (0.0) 0 | 1 (0.4) 1 | 1 (1.3) 1 |
| Arthralgia | 4 (1.7) 4 | 0 (0.0) 0 | 0 (0.0) 0 | 1 (1.3) 1 |
| Myalgia | 5 (2.1) 5 | 0 (0.0) 0 | 1 (0.4) 1 | 0 (0.0) 0 |
|  | CORBEVAX™ | PLACEBO | CORBEVAX™ | PLACEBO |
|  | N=234 | N=78 | N=234 | N=78 |
| Nervous system disorders | 9 (3.8) 10 | 4 (5.1) 4 | 8 (3.4) 9 | 2 (2.6) 2 |
| Headache | 8 (3.4) 8 | 4 (5.1) 4 | 8 (3.4) 8 | 2 (2.6) 2 |
| Insomnia | 0 (0.0) 0 | 0 (0.0) 0 | 1 (0.4) 1 | 0 (0.0) 0 |
| Somnolence | 2 (0.9) 2 | 0 (0.0) 0 | 0 (0.0) 0 | 0 (0.0) 0 |
| Respiratory, thoracic & mediastinal disorders | 0 (0.0) 0 | 0 (0.0) 0 | 2 (0.9) 2 | 0 (0.0) 0 |
| Nasopharyngitis | 0 (0.0) 0 | 0 (0.0) 0 | 1 (0.4) 1 | 0 (0.0) 0 |
| Rhinorea | 0 (0.0) 0 | 0 (0.0) 0 | 1 (0.4) 1 | 0 (0.0) 0 |
| Skin & Subcutaneous tissue disorders | 1 (0.4) 1 | 0 (0.0) 0 | 0 (0.0) 0 | 0 (0.0) 0 |
| Urticaria | 1 (0.4) 1 | 0 (0.0) 0 | 0 (0.0) 0 | 0 (0.0) 0 |

Percentages were calculated using column header count as denominator.
N<sub>1</sub>: Subject Count, N: Sample Size, n:Event Count
**General Note:** • All AE's were represented as: Subject count (Percentage of subjects) Event Count.

All the reported adverse events were mild to moderate in their intensity. There were no serious AEs/grade-3 AEs/deaths/AESIs reported during the study period and all the reported AEs resolved without any sequelae. Severity and causality of reported AEs were presented in table 4. Most of the AEs were related to vaccination. The safety profile of CORBEVAX™ arm in <18 to ≥5 years’ children and adolescents was found to be safe. The adverse event profile was also found to be acceptable.

**Table 4:** Overview of AEs by Severity & Causality.

| Category<br>N1 (%) n | <18 to ≥12 years (n=312) |  | <12 to ≥5 years (n=312) |  |
| --- | --- | --- | --- | --- |
|  | CORBEVAX™ | PLACEBO | CORBEVAX™ | PLACEBO |
|  | N=234 | N=78 | N=234 | N=78 |
| Number of subjects with at least one AE | 66 (28.2) 118 | 22 (28.2) 35 | 52 (22.2) 95 | 26 (33.3) 45 |
| Number of subjects with at least one SAE | 0 (0.0) 0 | 0 (0.0) 0 | 0 (0.0) 0 | 0 (0.0) 0 |
| Number of subjects with at least one MAAE | 1 (0.4) 2 | 0 (0.0) 0 | 0 (0.0) 0 | 0 (0.0) 0 |
| Severity |  |  |  |  |
| Mild | 65 (27.8) 116 | 21 (26.9) 34 | 51 (21.8) 94 | 26 (33.3) 45 |
| Moderate | 2 (0.9) 2 | 1 (1.3) 1 | 1 (0.4) 1 | 0 (0.0) 0 |
| Severe | 0 (0.0) 0 | 0 (0.0) 0 | 0 (0.0) 0 | 0 (0.0) 0 |
| Life Threatening | 0 (0.0) 0 | 0 (0.0) 0 | 0 (0.0) 0 | 0 (0.0) 0 |
| Causality |  |  |  |  |
| Very likely / Certain | 25 (10.7) 48 | 10 (12.8) 16 | 28 (12.0) 58 | 13 (16.7) 20 |
| Probable | 30 (12.8) 39 | 10 (12.8) 15 | 23 (9.8) 30 | 11 (14.1) 18 |
| Possible | 20 (8.5) 30 | 3 (3.8) 3 | 6 (2.6) 6 | 3 (3.8) 4 |
| Unlikely | 0 (0.0) 0 | 1 (1.3) 1 | 0 (0.0) 0 | 1 (1.3) 1 |
| Unrelated | 1 (0.4) 1 | 0 (0.0) 0 | 1 (0.4) 1 | 1 (1.3) 2 |
| Unclassifiable | 0 (0.0) 0 | 0 (0.0) 0 | 0 (0.0) 0 | 0 (0.0) 0 |
|  | CORBEVAX™ | PLACEBO | CORBEVAX™ | PLACEBO |
|  | N=234 | N=78 | N=234 | N=78 |
| Related (Certain/Probable/ Possible) | 66 (28.2) 117 | 21 (26.9) 34 | 52 (22.2) 94 | 25 (32.1) 42 |
| Unrelated (Unlikely/Unrelated/<br>Unclassifiable) | 1 (0.4) 1 | 1 (1.3) 1 | 1 (0.4) 1 | 2 (2.6) 3 |
| Number of subjects discontinued due to AE | 0 (0.0) 0 | 0 (0.0) 0 | 0 (0.0) 0 | 0 (0.0) 0 |

Percentages were calculated using column header count as denominator. N<sub>1</sub>: Subject Count, N: Sample Size, n: Event Count.
**General Note:** All AE's were represented as: Subject count (Percentage of subjects) Event Count.

### Immunogenicity Findings

#### Anti-RBD IgG and sub-class assessment in pediatric population in comparison to adult population

Anti-RBD IgG data is available at day0 and day42 in 229 subjects each in 12 to <18 Years’ and 5 to <12 Years’ age-cohorts who received CORBEVAX™ vaccine. The corresponding data for the adult population tested as part of earlier clinical trial, BECT-069-Phase-2/3 study^13^, is also provided for comparison. The data is summarized in table 5 and shown in figure 2. Seroconversion was observed in 91% and 96% of 12 to <18 Years’ and 5 to <12 years’ age-cohorts respectively, whereas seroconversion noted in 94% of subjects in adult population vaccinated with CORBEVAX™.

**Table 5:** Summary of Anti-RBD IgG concentration at Day-0 and Day-42 time points.

| Day-0 Testing<br>GMC; EU/mL | Day-42 Testing GMC;<br>EU/mL | Geometric Mean Fold<br>Rise post vaccination | % Seroconversion post-vaccination |
| --- | --- | --- | --- |
| **Age Group : 12 to <18 Years' n=229** |  |  |  |
| 939 | 18049 | 19 | 91% (209 out of 229) |
| **Age group 5 to <12 Years' n=229** |  |  |  |
| 964 | 26802 | 28 | 96% (220 out of 229) |
| **Age Group 18-55 Years' – from BECT-069 Phase II study, n=100** |  |  |  |
| 945 | 26448 | 28 | 94% |

**Figure 2.**
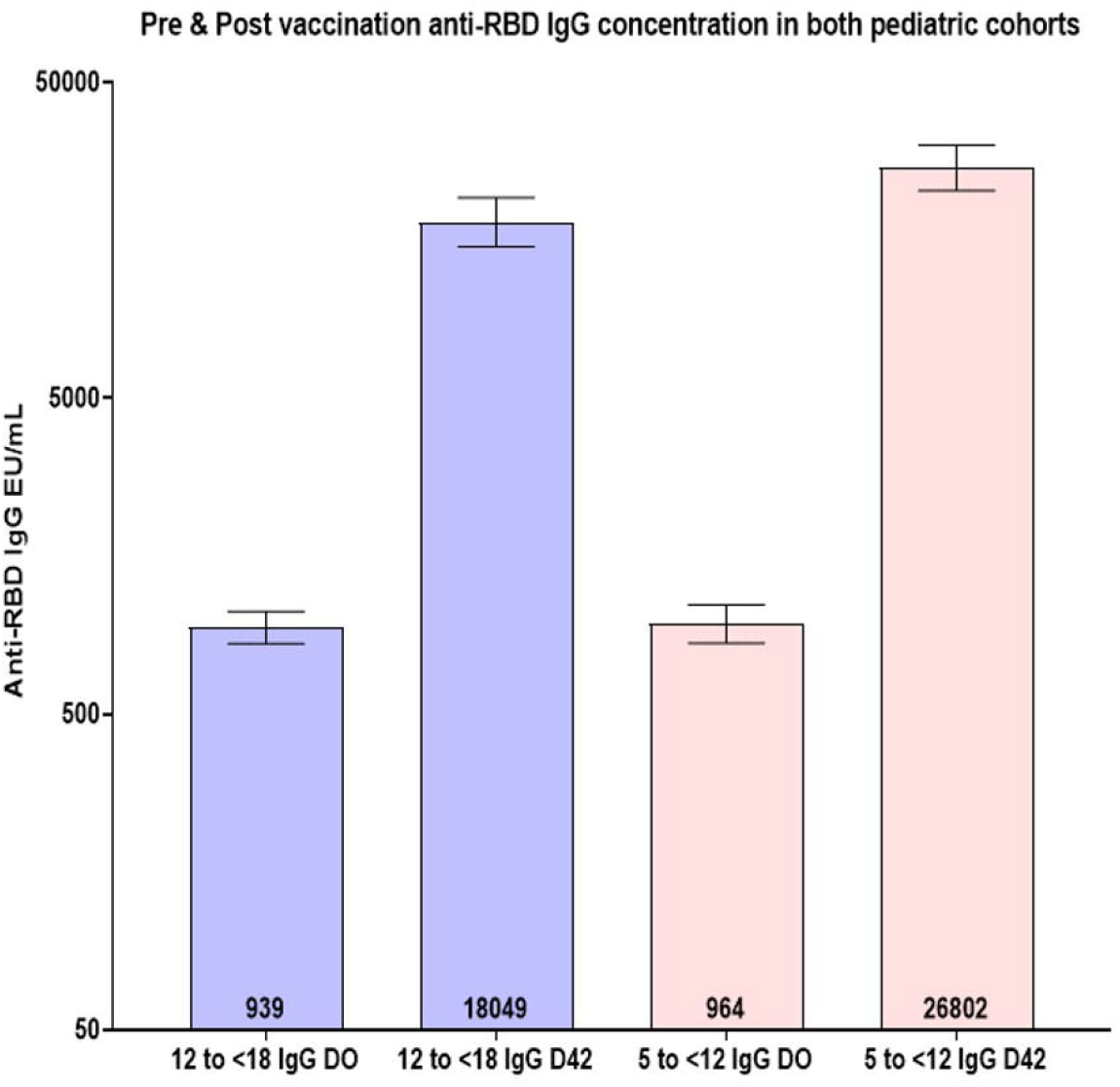
Anti-RBD lgG concentration in the two age-groups, pre (day-0) and post-vaccination (day-42) Summary of anti-RBD lgG concentrations in both pediatric age sub groups: A) Anti-RBD lgG concentrations were induced significantly high in CORBEVAX™ vaccinated subjects measured at day 42 in comparison to day 0. GMCT’s with 95% Confidence Interval (two-sided bars) are included in the figure.

The Th1 vs Th2 skew in the humoral immune response was assessed by measuring the anti-RBD titers for two IgG subclasses i.e. IgG1 and IgG4. The Geometric Mean Ratio of IgG1 to IgG4 titers post-vaccination was 40 and 45 fold in the 12 to <18 Years’ and 5 to <12 Years’ age-cohorts respectively indicating desired Th1 skew to the humoral immune response. The data is summarized in table 6 and graphically in figure 3 in comparison to adult population.

**Table 6:** Summary of Anti-RBD IgG1 & IgG4 titers at Day-0 and Day-42 time points.

| Day-0 |  | Day-42 |  |  |
| --- | --- | --- | --- | --- |
| IgG1<br>GMT | IgG4<br>GMT | IgG1<br>GMT | IgG4<br>GMT | Geometric Mean Ratio of<br>IgG1 to IgG4 Titers @ D42 |
| **Age Group : 12 to <18 Years' n=229** |  |  |  |  |
| 85 | 39 | 3494 | 87 | 40 |
| **Age Group : 5 to <12 Years' n=229** |  |  |  |  |
| 78 | 40 | 4654 | 108 | 43 |
| **Age Group 18-55 Years' – from BECT-069 Phase II study, n=100** |  |  |  |  |
| 126 | 41 | 7167 | 94 | 76 |

**Figure 3.**
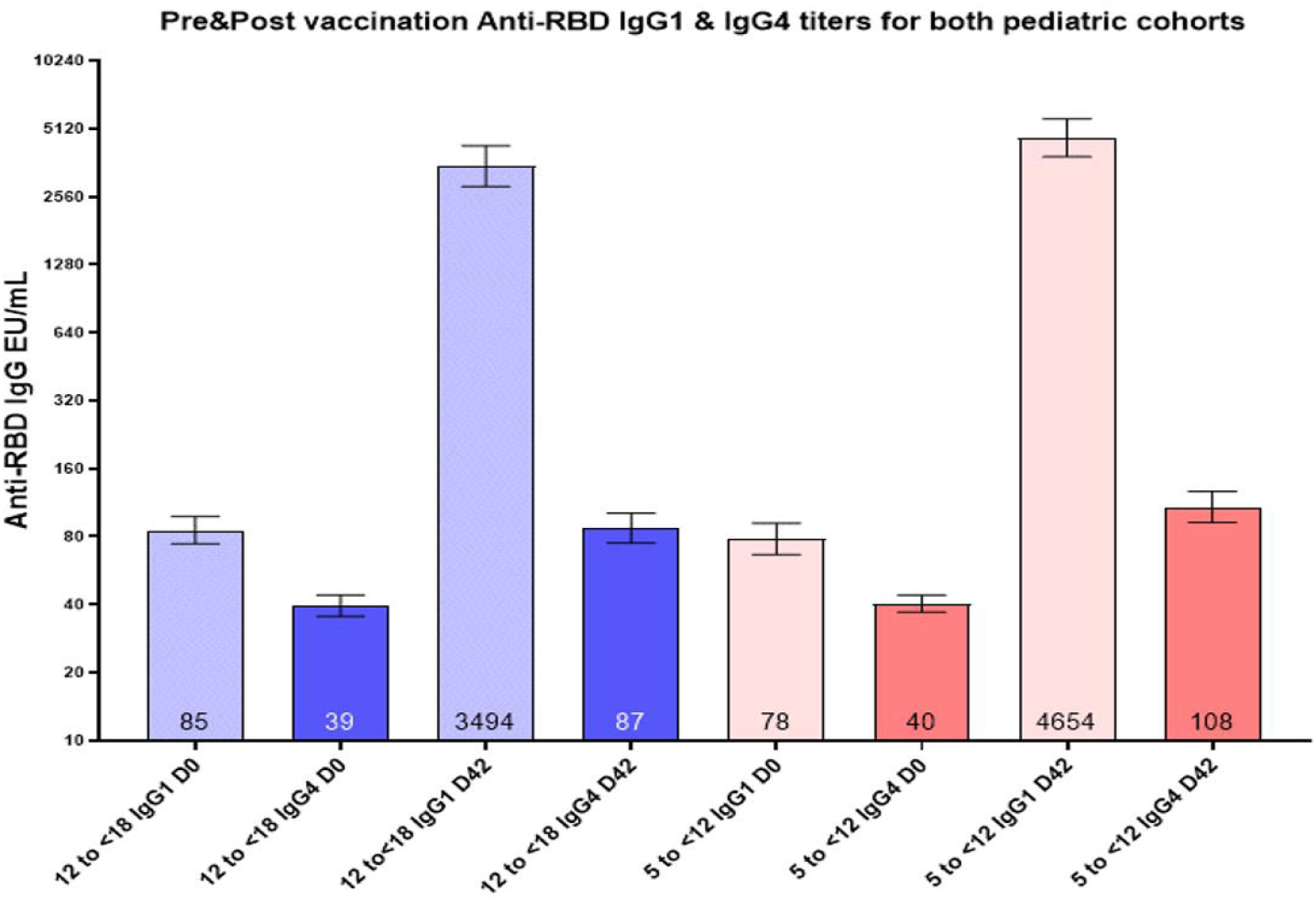
Anti-RBD lgG1 and lgG4 sub class titers in the two age-groups, pre (day-0) and post-vaccination (day-42) Summary of anti-RBD lgG sub class titers in both pediatric age sub groups: A) Anti-RBD lgG1 and lgG4 geometric mean concentrations were measured at day0 and day 42 post vaccination. In both age sub groups, lgG1 subclass titers predominantly increased after vaccination with CORBEVAX™ whereas lgG4 levels were comparable at day 0 and day 42 time points, indicating that CORBEVAX™ vaccine induces Th1 skewed immune responses. GMCT’s with 95% Confidence Interval (two-sided bars) are included in the figure.

#### Neutralizing Antibody Titers in pediatric population in comparison to adult population

Neutralizing Antibody titers (nAbs) were tested in the subject sera samples collected prior to vaccination (Day-0) and fourteen days after two doses of vaccination (Day-42) against Wild-Type SARS-COV-2 strains and Delta strain in a Micro Neutralization Assay. Interim data is available for 444 of 468 subjects (224 and 220 subjects in 12 to

<18 Years and 5 to <12 Years age-cohorts respectively) who received CORBEVAX™ vaccine is presented here. Data generated in pediatric population was compared with adult population from previous clinical trial as mentioned above. The data is summarized in table 7 and shown graphically in figure 4.

**Table 7:**
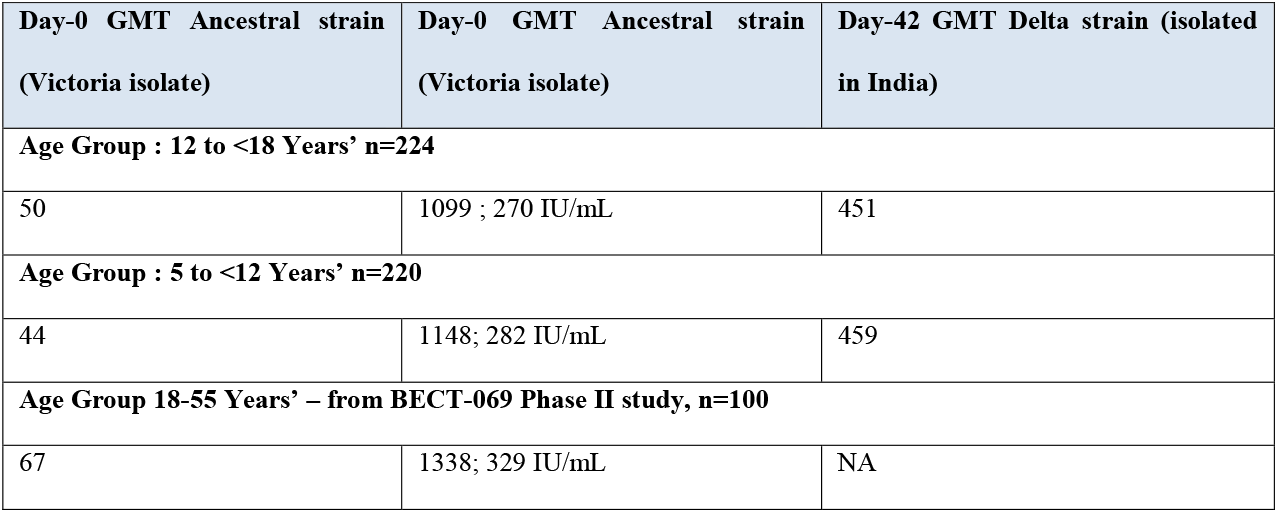
Summary of nAb titers pre and post vaccination in both age groups.

| Day-0 GMT Ancestral strain<br>(Victoria isolate) | Day-0 GMT Ancestral strain<br>(Victoria isolate) | Day-42 GMT Delta strain (isolated<br>in India) |
| --- | --- | --- |
| **Age Group : 12 to <18 Years' n=224** |  |  |
| 50 | 1099 ; 270 IU/mL | 451 |
| **Age Group : 5 to <12 Years' n=220** |  |  |
| 44 | 1148; 282 IU/mL | 459 |
| **Age Group 18-55 Years' – from BECT-069 Phase II study, n=100** |  |  |
| 67 | 1338; 329 IU/mL | NA |

**Figure 4.**
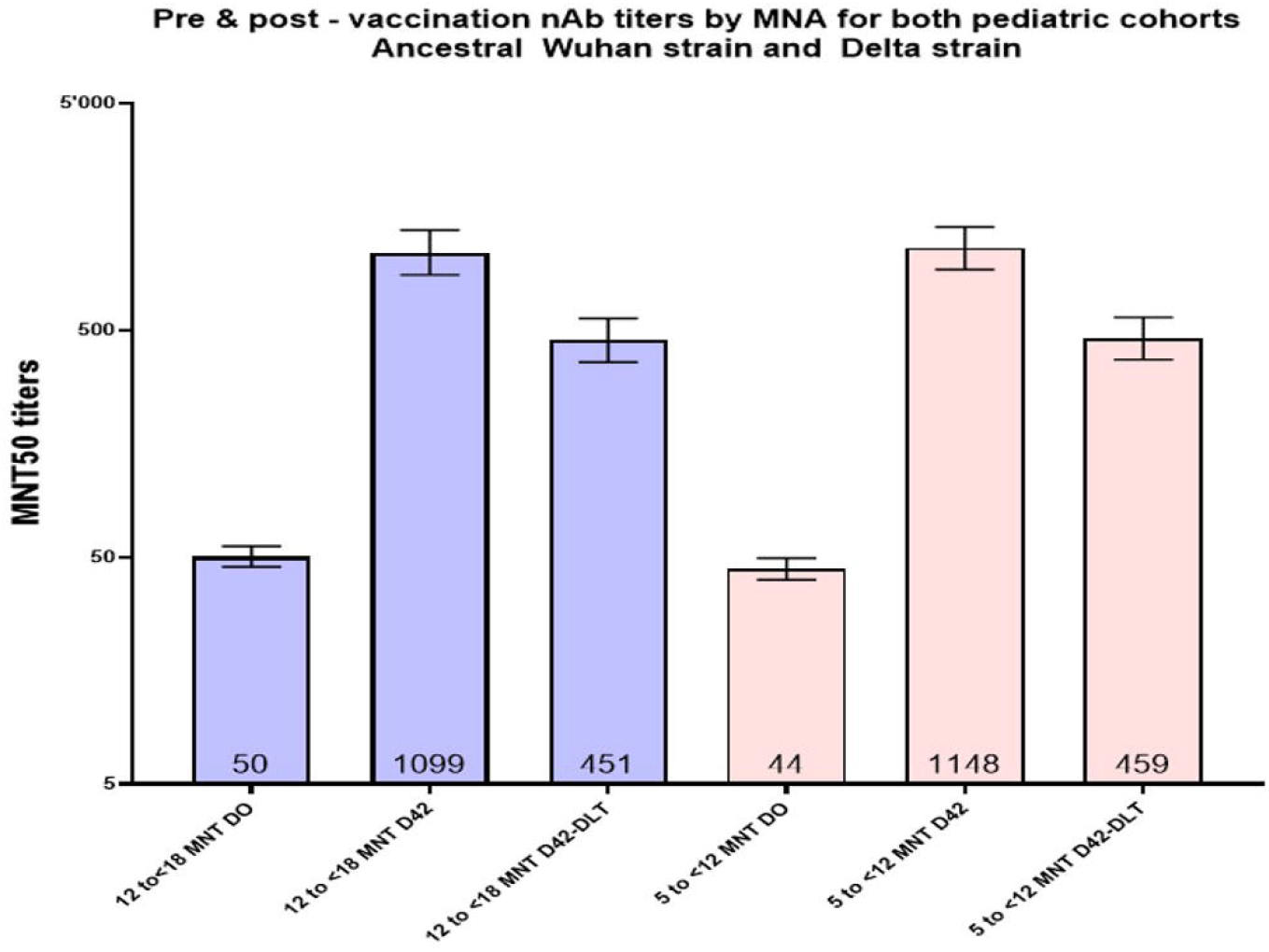
Neutralizing antibody titers against Ancestral and Delta strains of SARS-COV-2. **nAb titers measured against ancestral and Delta strains of SARS-COV-2:** Neutralizing Antibody titers were tested in the subject sera samples collected prior to vaccination (Day-0) and fourteen days after two doses of vaccination (Day-42). The testing was conducted against Wild-Type SARS-COV-2 strain and Delta strain in a Micro Neutralization Assay **(MNA)**. The observed nAb GMT’s in both age groups that received CORBEVAX™ vaccine are much higher than the thresholds induced by other COVID-19 vaccines, which is indicative of >90% vaccine effectiveness of CORBEVAX™ vaccine. GMCT’s with 95% Confidence Interval (two-sided bars) are included in the figure.

The Day-42 nAb GMT’s in pediatric population against the ancestral Wuhan strain are significantly higher than two key thresholds that are indicators of high vaccine effectiveness.

1. Comparison with Human Convalescent Sera Panel GMT: PHE, UK conducted testing of convalescent sera samples obtained from 32 RT-PCR positive COVID-19 patients with severe disease and the reported GMT from this panel was 522. As per the publication by Khoury et al. (Neutralizing antibody levels are highly predictive of immune protection from symptomatic SARS-CoV-2 infection,^14^ GMT ratio of >1.5 for vaccinated sera to HCS panel is indicative of very high vaccine effectiveness in terms of prevention of symptomatic infection
2. Comparison with Correlates of Protection information: nAb titers vs. symptomatic infection assessment conducted as part of Moderna mRNA-1273 vaccine (Spikevax) and AstraZeneca AZD1222 vaccine (Vaxzevria) Phase III efficacy trials showed that post-vaccination nAb GMT’s of 100 IU/mL and 60 IU/mL are indicative of vaccine efficacy of >90% and >80% respectively. The observed nAb GMT’s in both age groups that received CORBEVAX™ are much higher than these thresholds which is indicative of >90% vaccine effectiveness

#### Non-Inferiority comparison between pediatric and adult population nAb titers

The nAb Geometric Mean titers were calculated for both the age-groups and the ratio of each cohort GMT to adult GMT was calculated. Then the variance of each cohort was calculated from the log transformed data and then the combined variance of the two comparator populations was determined. The lower bound of the 95% confidence interval of the ratio of the two GMT’s was then determined by subtracting the variance multiplied by the correction factor from the ratio of the two GMT’s. The non-inferiority calculation was summarized in table 8. For both the pediatric age cohorts, the lower bound of the ratio of the GMT with adult cohort was >0.5 which as the threshold for demonstration of non-inferiority. The Geometric Mean Fold Rise (GMFR) in nAb titer from baseline to Day-42 time-point are 22 and 26 respectively for 12 to <18 and 5 to <12 cohorts respectively. These pediatric GMFR’s are higher than the adult cohort GMFR of 20.

**Table 8:**
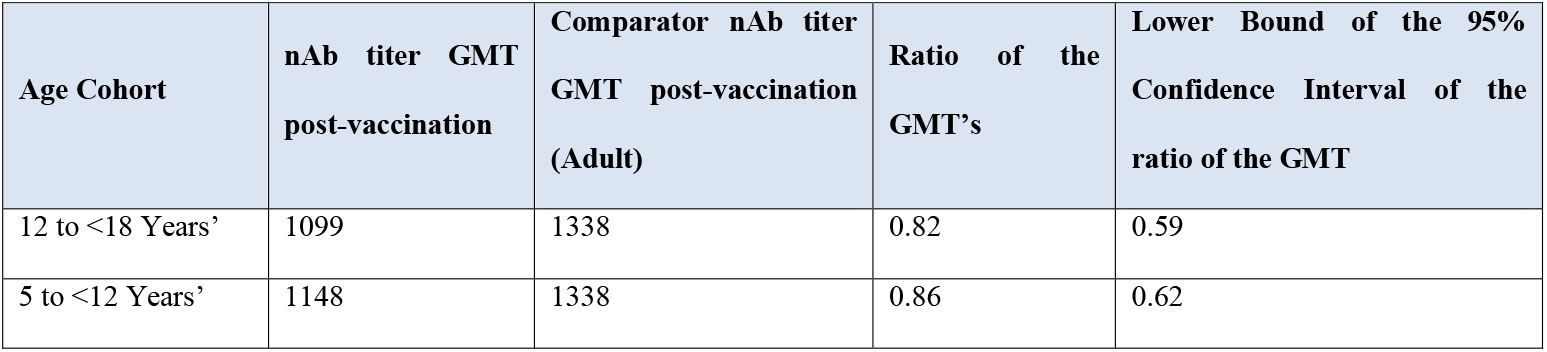
Non-inferiority assessment of the nAb titers of pediatric vs adult population.

#### Cellular Immune Response Assessment

Cellular immune response involved direct measurement of cytokines secreted by stimulation of whole blood samples with SARS-COV-2 peptides using TrueCulture technology. Table 9 summarizes the average concentrations of Interferon-gamma and Interleukin-4 cytokines recorded at Day-0 and Day-42 time-points from 18 subjects in the 12 to <18 years’ age-group that received CORBEVAX™ vaccine. Substantial increase was observed in the IFN-gamma concentration at Day-42 time-point under activated conditions in comparison with IL-4 concentration. This indicates significant and Th1 skewed cellular immune response generated from CORBEVAX™ vaccination

**Table 9:** Summary of cytokine concentrations at Day-0 and Day-42 time-points-TrueCulture technology.

| Cytokine; average concentration | Day-0 Time point |  | Day-42 Time-point |  |
| --- | --- | --- | --- | --- |
|  | Null | Activated | Null | Activated |
| **Pediatric cohort : 5 to <18 Years; N=18** |  |  |  |  |
| **Interferon-gamma; pg/mL** | 3.38 | 9.03 | 5.99 | 69.47 |
| **Interleukin-4; pg/mL** | 0.61 | 0.85 | 0.98 | 0.98 |
| **Adult cohort : 18 to 55 Years (from BECT-069-Phase II study) ; N=67** |  |  |  |  |
| **Interferon-gamma; pg/mL** | 7.03 | 26.24 | 2.63 | 99.82 |
| **Interleukin-4; pg/mL** | 5.19 | 4.26 | 8.10 | 10.96 |

The total Interferon-gamma secreting PBMC’s were measured upon stimulation with SARS-COV-2 RBD peptides were measured for seven subjects in the pediatric population at this interim stage using ELISPOT method. ELISPOT analysis was not conducted as part of earlier adult BECT-069-Phase II study. Hence for comparison purpose the data generated in adult population in BECT-074-Phase III study was used (manuscript under preparation). The comparison is shown in figure 5 and data was summarized in table 10. The median spot forming units were 88 and 120 in pediatric and adult population respectively, re-emphasizing the fact that CORBEVAX™ vaccine skewing immune responses towards desired Th1 type in both pediatric and adult populations.

**Figure 5.**
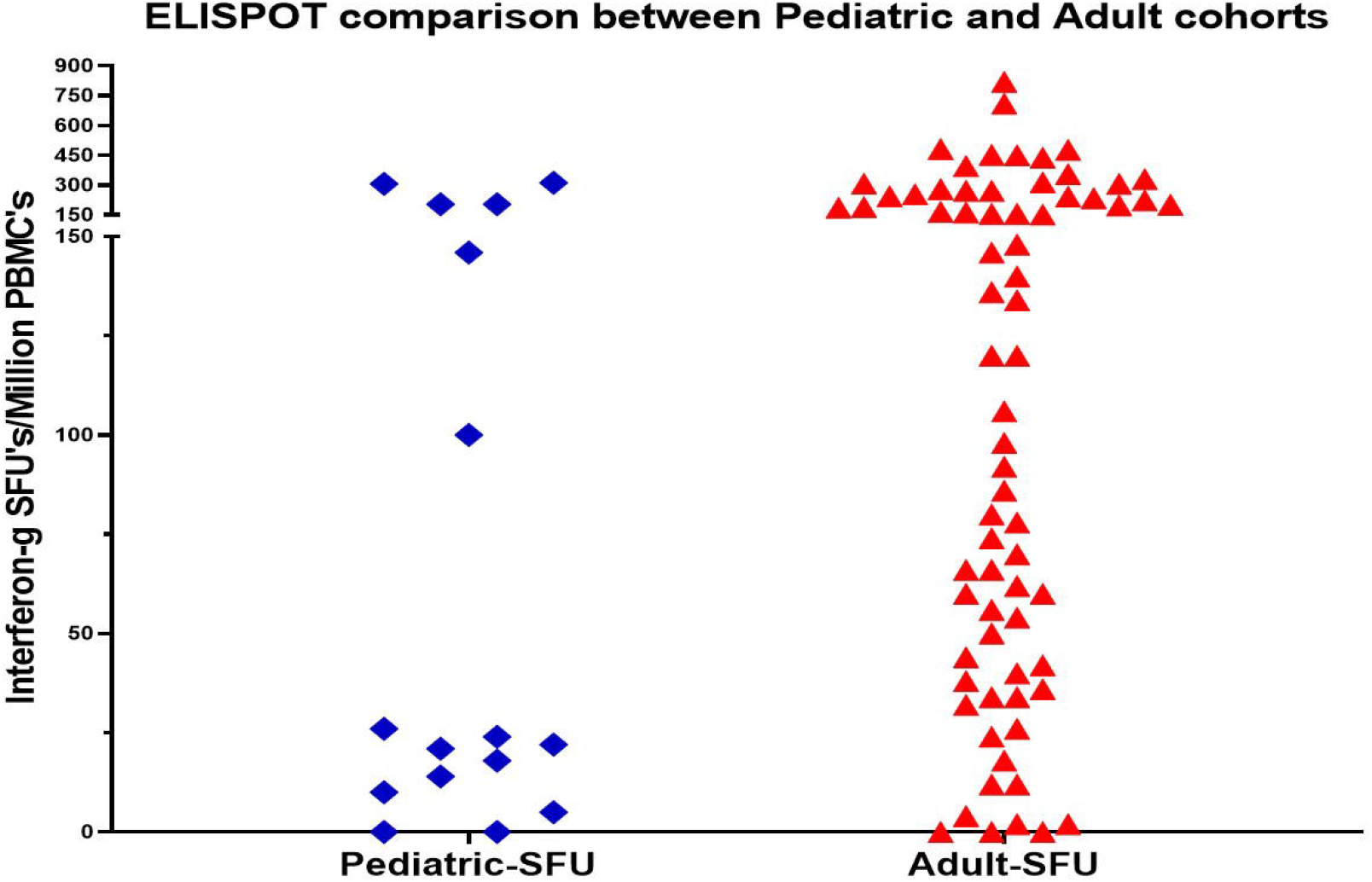
Cellular immune response assessment induced by CORBEVAX™ vaccine in pediatric population. **Comparison of cellular immune response in pediatric and adult population via ELISPOT:** The total Interferon-gamma secreting PBMC’s upon stimulation with SARS-COV-2 RBD peptides were measured for sixteen subjects in the pediatric population. The magnitude of cellular immune response in the pediatric cohort was similar to the adult population in terms of Interferon-gamma ELISPOT SFU’s

**Table 10:** Comparison of cellular immune (IFN-gamma) response via ELISPOT in pediatric and adult population.

| Age Cohort | Total subject samples evaluated<br>post vaccination | Average SFU | Median SFU (IQR) |
| --- | --- | --- | --- |
| 5 to <18 Years' | 16 | 88 | 23 (11 – 190) |
| 18 to 80 Years' | 73 | 176 | 120 (43 – 249) |

## DISCUSSION

In this trial, safety and immunogenicity of a sub-unit protein vaccine for Covid-19, CORBEVAX™ was studied in children (5-11 years) and Adolescents (12-17 years). Results indicated that CORBEVAX™ is safe and well tolerated with no vaccine related serious adverse events, MAAEs or AESI when administered to healthy children and adolescents. Consistent humoral immune response post-CORBEVAX™ vaccination was observed in both the pediatric age cohorts in terms of anti-RBD IgG concentrations, percentage seroconversion and neutralizing antibody (nAb) titers. Significant nAb titers were observed against both ancestral Wuhan and Delta strains which has also been consistently noted in all clinical trials of CORBEVAX™ conducted so far. The nAb titers post CORBEVAX™ vaccination are indicative of very high vaccine effectiveness when compared with the established HCS panel threshold or Correlate of Protection observed in vaccine efficacy trials. The nAb titers observed in both the pediatric age cohorts were non-inferior to the adult cohort in terms of ratio of the GMT’s of both the cohorts. The humoral immune response was skewed towards Th1 category as indicated by the high ratio of anti-RBD IgG1 titers vs. IgG4 titers. The magnitude of cellular immune response in the pediatric cohort was similar to the adult population^13^ in terms of ELISPOT SFU comparisons. The cellular immune response in the pediatric population also demonstrated the expected Th1 skew as indicated by significant IFN-γ expression by the stimulated PBMCs and minimal expression of IL-4 cytokine; as was observed in adult population.

Approved vaccines in adolescents are effective with almost 100% efficacy. However, safety issues were reported across several studies. Messenger RNA COVID-19 vaccine BNT162b2 is one of the most widely used vaccines in children and adolescents aged as young as 5 years in several countries.^15^ Nevertheless, a population-based retrospective cohort study and a large cohort study of nearly a million individuals and other studies have reported adverse events of special interest (myocarditis and sleeping disturbance) following the second dose of BNT162b2 in adolescents.^16-21^ Similar AESI were observed in adolescents receiving another mRNA (Moderna) vaccine.^22^

CORBEVAX™ was found to be safe without any observed severe, serious adverse events, AESI and MAAEs in both children and adolescents post second dose. Moreover, CORBEVAX™ uses a traditional recombinant protein-based technology (which is also used for the production of vaccines like Hepatitis B) that has a proven safety record in children making the vaccine more reliable, cost effective and safe to be administered. In addition, this platform enables the production of vaccine at large scales making it widely accessible to inoculate the global population.

### Study limitations

Limitations of the current study includes: Long-term safety in pediatric population was not established yet as interim results are available only until day 56. Safety profile of the CORBEVAX™ vaccine will be established up to 6 months after second dose of the vaccination. Immunogenicity of the candidate vaccine was studied but the efficacy of the vaccine against Covid-19 infection was not studied. Also, there is a need for evaluating antibody persistence in longer follow-up period and neutralizing potential against new Omicron variant was not tested in the current study which will be addressed in the future studies.

In conclusion, CORBEVAX™ vaccine is safe, well tolerated and elicited excellent antibody and cellular immune responses that can offer significant protection against symptomatic infection from SARS-CoV-2 virus in pediatric population aged 5-17 years.

## Data Availability

Additional study data which is not part of the manuscript can be made available upon request and addressed to the corresponding author Dr. Subhash Thuluva at his

## Notes

**Funding Support:** The study was funded by grants from BIRAC- a division of the Department of Biotechnology, Govt of India.

**Conflicts of Interest:** ST, VP, SG, VY, RM, PVS, KT, MK, SKM, SA and ASJ are employees of Biological E Limited and they don’t have any incentives or stock options. All other participating authors declare no competing interests.

### Competing Interest Statement

The authors have declared no competing interest.

### Clinical Trial

CTRI/2021/10/037066

### Author Declarations

The Investigational Review Board or Ethics Committee at each study site approved the protocol. All participants provided written informed consent before enrollment into the study. 1IEC-Mysore Medical College and Research Institute, Irwin Road, Mysuru 57000, Karnataka, IndiaApproved 08-10-2021ECR/134/Inst/KA/2013/RR-19 2Institutional Ethics Committee JSS Medical College, JSS Medical College and Hospital, Sri Shivarathreeshwara Nagar, Mysuru - 570015, Karnataka, India Approved 05-10-2021ECR/387/Inst/KA/2013/RR-19 3Ethics Committee Guru Teg Bahadur Hospital Dilshad Garden, Delhi - 110095 IndiaApproved 06-10-2021ECR/510/Inst/DL/2014/RR-20 4Institutional Ethics Committee, KLE University KLE University KLE Dr.PK Hospital and MRC, Nehru Nagar, Belagavi 590010 Karnataka, India. Approved 26-10-2021ECR/211/Inst/KA/2013/RR-19 5Shubham Sudbhawana Superspeciality Hospital - Ethics Committee, B 31/80, 23B - Bhogabeer, Lanka, Varanasi - 221005, Uttar Pradesh, IndiaApproved 09-10-2021ECR/667/Inst/UP/2014/RR-20 6Institutional Ethics Committee, Government Medical College and Hospital, Government Medical College Medical Square, Hanuman Nagar Nagpur 440003, Maharashtra, IndiaApproved 25-10-2021ECR/43/Inst/MH/2013/RR-19 7IEC NIMS University, Hotam administrative block, Delhi- Jaipur highway, Jaipur- 303121, Rajasthan, India. Approved 04-10-2021ECR/665/Inst/RJ/2014/RR-17 8Sant Dnyaneshwar Medical Education & Research Centre - Institutional Review Board, 695/A, Sadashiv Peth, Opp. Vijay Talkies, Laxmi Road, Pune 411030, MaharashtraApproved 27-10-2021ECR/127/Inst/MH/2013/RR-19 9ETHICS COMMITTEE, St. Theresa's Hospital, Sanath Nagar, Opp. Erragada Raitu Bazar, Hyderabad 500018, Telangana, IndiaApproved 01-10-2021ECR/230/Inst/AP/2013/RR-19 10IEC, Mahatma Gandi Institute Of Medical Sciences, Sewagram, Wardha 442102, Maharshtra, india Approved 23-10-2021ECR/47/Inst/MH/2013/RR-19 11Ethics Committee Dy.Patil Vidyapeeth, Pimpri, Pune - 411018, Maharashtra, IndiaApproved 07-12-2021ECR/361/Inst/MH/2013/RR-19 12IRB Christian Medical college, Office of Research, 1st floor, Carman Block, Bagayam, Vellore 632002, Tamilnadu, India. Approved 30-10-2021ECR/RENEW/INST/2019/3526 13Institutional Ethics Committee for ESIC Medical College And Hospital NH-3, NIT, Behind BK Hospital, Faridabad 121001, Haryana, IndiaApproved 16-10-2021ECR/1539/Inst/HR/2021 14IEC Prakhar Hospital Pvt Ltd, 8/219, Arya Nagar, Kanpur 208002, Uttar Pradesh, India. Approved 20-10-2021ECR/1017/Inst/UP/2017/RR-21 15Aakash Healthcare Institutional Ethics Committee, Road No. 201, Sector 3, Dwarka, New Delhi - 110075, IndiaApproved 09-11-2021ECR/1265/Inst/DL/2019 16Gullurkar Hospital Ethics Committee, Plot no. 20, Reshimbag Umred Road, Nagpur- 440009, Maharashtra, IndiaApproved 18-11-2021ECR/1374/Inst/MH/2020 17Institutional Ethics Committee- Induss Hospital, 13-23-93/1, Krishnaveninagar, Gaddiannaram, Near Muncipal Office, Saroornagar, Ranga Reddy Dist., Hyderabad 500035, Telangana, IndiaApproved 06-11-2021ECR/1606/Inst/TG/2021 18Institutional Ethics Committee- Jeevan Rekha Hospital, Dr. B.R. Ambedkar Road, Opp Civil Hospital, Belagavi- 590002, Karnataka, IndiaApproved 06-11-2021ECR/1242/Inst/KA/2019 19Institutional Ethics Committee - Jawahar Lal Nehru Medical College, Kala Bagh Ajmer - 305001, Rajasthan, IndiaApproved 22-11-2021ECR/1156/Inst/RJ/2018/RR-22 20Lifepoint Research- Ethics Committee, 145/1, Mumbai-Bangalore highway, Near Hotel Sayaji, Wakad, Pune - 411057, Maharashtra, IndiaApproved 09-11-2021ECR/751/Inst/MH/2015/RR-21 21Penta-Med Ethics Committee, Medipoint Hospitals Pvt. Ltd, 241/1, New D.P.Road, Near Sai Heritage, Pune - 411007, Maharashtra, IndiaApproved 12-11-2021ECR/357/Inst/MH/2013/RR-20 22Unique Children's Hospital Ethics Committee, Hira Moti Fortune Opp. Police station, Pune Mumbai Road, Chinchwad, Pune-411019, Maharashtra, IndiaApproved 16-12-2021ECR/1203/Inst/MH/2019 23CARE-IHEC for Faculty Research Chettinad Academy Of Research And Education, Rajiv Gandhi Salai (OMR),Kelambakkam, Kanchipuram - 603103, Tamil Nadu, IndiaApproved 09-12-2021ECR/1589/Inst/TN/2021

## REFERENCES

1. Feldstein LR, Tenforde MW, Friedman KG, et al. Characteristics and Outcomes of US Children and Adolescents With Multisystem Inflammatory Syndrome in Children (MIS-C) Compared With Severe Acute COVID-19. JAMA 2021; 325(11): 1074–87.

2. Hageman JR. Long COVID-19 or Post-Acute Sequelae of SARS-CoV-2 Infection in Children, Adolescents, and Young Adults. Pediatr Ann 2021; 50(6): e232–e3.

3. Sperotto F, Friedman KG, Son MBF, VanderPluym CJ, Newburger JW, Dionne Cardiac manifestations in SARS-CoV-2-associated multisystem inflammatory syndrome in children: a comprehensive review and proposed clinical approach. Eur J Pediatr 2021; 180(2): 307–22.

4. Seth S, Rashid F, Khera K. An overview of the COVID-19 complications in paediatric population: A pandemic dilemma. Int J Clin Pract 2021; 75(9): e14494.

5. Chu VT, Yousaf AR, Chang K, et al. Household Transmission of SARS-CoV-2 from Children and Adolescents. N Engl J Med 2021; 385(10): 954–6.

6. Yang J, Zhang T, Qi W, et al. COVID-19 vaccination in Chinese children: a cross-sectional study on the cognition, psychological anxiety state and the willingness toward vaccination. Hum Vaccin Immunother 2022; 18(1): 1–7.

7. Ishimoto Y, Yamane T, Matsumoto Y, Takizawa Y, Kobayashi K. The impact of gender differences, school adjustment, social interactions, and social activities on emotional and behavioral reactions to the COVID-19 pandemic among Japanese school children. SSM Ment Health 2022; 2: 100077.

8. Frenck RW, Jr., Klein NP, Kitchin N, et al. Safety, Immunogenicity, and Efficacy of the BNT162b2 Covid-19 Vaccine in Adolescents. N Engl J Med 2021; 385(3): 239–50.

9. Ali K, Berman G, Zhou H, et al. Evaluation of mRNA-1273 SARS-CoV-2 Vaccine in Adolescents. N Engl J Med 2021; 385(24): 2241–51.

10. Han B, Song Y, Li C, et al. Safety, tolerability, and immunogenicity of an inactivated SARS-CoV-2 vaccine (CoronaVac) in healthy children and adolescents: a double-blind, randomised, controlled, phase 1/2 clinical trial. Lancet Infect Dis 2021; 21(12): 1645–53.

11. Xia S, Zhang Y, Wang Y, et al. Safety and immunogenicity of an inactivated COVID-19 vaccine, BBIBP-CorV, in people younger than 18 years: a randomised, double-blind, controlled, phase 1/2 trial. Lancet Infect Dis 2022; 22(2): 196–208.

12. Open label phase I/II clinical trial and predicted efficacy of SARS-CoV-2 RBD protein vaccines SOBERANA 02 and SOBERANA Plus in children. MedRxiv 2022.

13. Thuluva S, Paradkar V, Turaga K, et al. Selection of optimum formulation of RBD-based protein sub-unit covid19 vaccine (Corbevax) based on safety and immunogenicity in an open-label, randomized Phase-1 and 2 clinical studies. MedRxiv 2022.

14. Khoury DS, Cromer D, Reynaldi A, et al. Neutralizing antibody levels are highly predictive of immune protection from symptomatic SARS-CoV-2 infection. Nat Med 2021; 27(7): 1205–11.

15. Interim recommendations for use of the Pfizer–BioNTech COVID-19 vaccine, BNT162b2, under Emergency Use Listing.

16. Shay DK, Shimabukuro TT, DeStefano F. Myocarditis Occurring After Immunization With mRNA-Based COVID-19 Vaccines. JAMA Cardiol 2021; 6(10): 1115–7.

17. Mevorach D, Anis E, Cedar N, et al. Myocarditis after BNT162b2 mRNA Vaccine against Covid-19 in Israel. N Engl J Med 2021; 385(23): 2140–9.

18. Barda N, Dagan N, Ben-Shlomo Y, et al. Safety of the BNT162b2 mRNA Covid-19 Vaccine in a Nationwide Setting. N Engl J Med 2021; 385(12): 1078–90.

19. Bozkurt B, Kamat I, Hotez PJ. Myocarditis With COVID-19 mRNA Vaccines. Circulation 2021; 144(6): 471–84.

20. Patone M, Mei XW, Handunnetthi L, et al. Risks of myocarditis, pericarditis, and cardiac arrhythmias associated with COVID-19 vaccination or SARS-CoV-2 infection. Nat Med 2022; 28(2): 410–22.

21. Witberg G, Barda N, Hoss S, et al. Myocarditis after Covid-19 Vaccination in a Large Health Care Organization. N Engl J Med 2021; 385(23): 2132–9.

22. Husby A, Hansen JV, Fosbol E, et al. SARS-CoV-2 vaccination and myocarditis or myopericarditis: population based cohort study. BMJ 2021; 375: e068665.

