## Supplementary Information for "Safety, tolerability and immunogenicity of Biological E’s CORBEVAX™ vaccine in children and adolescents: A Prospective, Randomised, Double-blind, Placebo controlled, Phase-2/3 Study"

**Supplementary Material**

Subject Eligibility

Only those subjects who fulfil ALL the inclusion and exclusion criteria will be enrolled into the study.

### **Inclusion Criteria**

Deviations from inclusion and exclusion criteria are not allowed because they can potentially jeopardize the scientific integrity of the study, regulatory acceptability or subject safety. Therefore, adherence to the criteria as specified in the protocol is essential.

Each subject must satisfy ALL of the following criteria at study entry:

1. Ability to provide written informed consent (by the parents or legally acceptable/authorized representative (LAR) and assent by the children aged between ≥7 to <18 years.
2. Subject, in the opinion of the investigator, has ability to communicate and willingness to comply with the requirements of the protocol.
3. Participants of either gender of age between <18years to ≥5 (Participant should be <18 years at the time of Screening of the study).
4. Participants virologically seronegative to SARS-CoV-2 infection by RT-PCR and anti-SARS-CoV-2 antibody prior to enrolment.
5. Participants considered of stable health as judged by the investigator, determined by medical history and physical examination with normal vital signs as defined in the protocol. *[Normal vital sign defined as body temperature <100.4ºF prior to enrolment].*
6. Agrees not to participate in another clinical trial at any time during the total study period.
7. Agrees to remain in the town where the study centre is located, for the entire duration of the study.
8. Willing to allow storage and future use of collected biological samples for future research in an anonymised form.

### **Exclusion Criteria**

The following criteria should be checked at the time of study entry. If ANY exclusion criterion applies, the subject must not be included in the study:

1. History of vaccination with any investigational vaccine against COVID-19 disease;
2. Seropositive to IgG antibodies against SARS CoV-2
3. Living in the same household of any COVID-19 positive person;
4. Use of any investigational or non-registered product other than the study vaccine during the trial period or 3 months prior to enrolment;
5. History of receipt of any licensed vaccine within 1 month prior to screening, likely to impact on interpretation of the trial data (e.g., influenza vaccines);
6. Current or planned participation in prophylactic drug trials for the duration of the study.
7. Known history of HIV 1 & 2, HBV and HCV infection in participants prior to enrolment.;
8. Body temperature of ≥100.4°F (>38.0°C) or symptoms of an acute illness at the time of screening or prior to vaccination;
9. History of severe psychiatric conditions likely to affect participation in the study;
10. History of any bleeding disorder (e.g. factor deficiency, coagulopathy or platelet disorder);
11. History of allergic disease or reactions likely to be exacerbated by any component of the Biological E’s CORBEVAX vaccine formulations;
12. Chronic respiratory disease, including asthma;
13. Chronic cardiovascular disease, gastrointestinal disease, liver disease, renal disease, endocrine disorder and neurological illness;
14. Any other serious chronic illness requiring hospital specialist supervision;
15. Chronic administration (defined as more than 14 days in total) of immunosuppressant (e.g. corticosteroids, cytotoxic drugs or antimetabolites, etc.) or other immune-modifying drugs (e.g. interferons) during the period starting six months prior to the first vaccine dose including use of any blood products. For corticosteroids, this will mean prednisolone ≥0.5 mg/kg/day, or equivalent. Inhaled and topical steroids are allowed;
16. Any confirmed or suspected immunosuppressive or immunodeficient condition, based on medical history and physical examination (no laboratory testing required);
17. Any medical condition that in the judgment of the investigator would make study participation unsafe.
18. Individuals who are part of the study team or close family members of individuals conducting the study.
19. Anaphylactic reaction following administration of the investigational vaccine.

**Methodology**

As per the kit manufacturer, subjects with antibody concentrations below 12 Antibody Units/mL were designated as sero-negative and were selected in the trial (Diasorin kit). Health status assessed during the screening period was based on medical history and clinical laboratory findings, vital signs, and physical examination.

**Procedure**

**Assessment of Reporting of Adverse events**

The period of observation for adverse events starts from the time the subject receives the single vaccination dose. All study subjects were observed at site for at least 60 minutes after vaccination for evidence of immediate reactions and in particular for symptoms of allergic phenomena (such as rashes or other allergic manifestations). Each subject’s parent / LAR was instructed to complete a diary card for 7 consequent days (Day 0 to Day 6) following first dose of vaccination, to record local and systemic symptoms. The diary was handed over to the PI at visit 3. Another diary was issued after 2^nd^ dose of vaccination at Day 28 and the subject’s parent / LAR was instructed to complete a diary card for 7 consequent days and for any other AE. All Serious Adverse Events (SAEs) and medically attended AEs if any, were collected and documented until the end of the study.

All adverse events occurred within the study period, regardless of severity, were monitored and medically managed by the investigator until resolution. All subjects experiencing adverse events - whether considered associated with the use of the study vaccine or not were monitored until symptoms subside and any clinically significant abnormal laboratory values if identified, have returned to baseline, or until there is a satisfactory explanation for the changes observed, or until death, in which case a full pathologist’s report should be supplied, if possible. All findings were reported on an “Adverse Events” eCRF page and on the “Serious Adverse Event” form as applicable, where necessary. All findings in subjects experiencing adverse events were reported also in the subject's medical records.

If any serious adverse event (SAE) occurred after the single dose vaccination until study end were reported to the Sponsor, Institutional Ethics committee (EC) and the licencing authority within 24 hrs of the occurrence of the SAE. The Sponsor and investigator after due analysis are responsible for the reporting of any unexpected or serious AEs to the Ethics committee (EC), Head of the Institute and the licencing authority within 14 days. All adverse events were followed up until resolution or until the end of the study whichever is earlier. The site investigator assessed the severity and causality of the event.

Safety and data monitoring wias carried out as per current ICH GCP, Indian GCP guidelines and G.S.R. 227(E), as applicable. The principal investigator reported/ will report safety issues to IEC and the sponsor periodically and as per regulatory requirement, until follow up visit of last subject.

**Immunogenicity Analysis**

Humoral immune responses were evaluated by following methods:

1. Anti-RBD Antibody response: Anti-RBD IgG concentration in the subject sera samples were measured at pre-vaccination (Day-0), post first dose (Day-28) and at multiple time-points post second dose (Day-42, Day-56, Day-208) using a validated enzyme linked immunosorbent assay (ELISA) method executed at Dang’s Lab, New Delhi, India. A monoclonal antibody, CR-3022 supplied by Lake Pharma Inc, CA, USA, that binds specifically to the RBD protein was used to generate standard curve of ELISA OD response vs. antibody concentrations. ELISA concentration equivalent to 1 ng/mL of CR-3022 antibody binding concentration was assigned concentration of 1 anti-RBD ELISA Unit/mL and thus anti-RBD IgG concentration in the sera samples were reported in EU/mL. The National Institute for Biological Standards and Control, UK plasma reference standard 20/130 was used as a positive control on all the plates with a control range of (**8151** to **15137** EU/mL) for validity consideration^1^. Geometric means were calculated for various subject cohorts at specific time-points and fold rise in anti-RBD concentrations for all time-points post start of vaccination were calculated in relation to the pre-vaccination concentrations and then geometric mean fold rise (GMFR) were calculated for each cohort.
2. Anti-RBD IgG subclass response: Anti-RBD IgG1 and IgG4 subclass titers were measured in the sera samples at pre-vaccination (Day-0), post first dose (Day-28) and at multiple time-points post second dose (Day-42, Day-56) using a validated ELISA method executed at Dang’s Lab, New Delhi, India. A pool of sera was prepared with very low anti-RBD IgG concentration from pre-vaccination samples and the threshold ELISA OD for assignment of titer was calculated as threshold OD = Average of blank pool OD + 5.56*standard deviation. Each sera sample was tested as a series of two-fold dilutions (starting from 50-Fold) and the dilution that resulted in OD higher than the threshold OD was assigned as the IgG1 or IgG4 titer. Geometric means were calculated for various subject cohorts at specific time-points and fold rise and GMFR in anti-RBD IgG1 and IgG4 titers for all time-points post vaccination were calculated in relation to the pre-vaccination titers.
3. SARS-COV-2 Virus Neutralization: SARS-COV-2 neutralizing antibody titers (nAb titers) were measured via 2 different methods^2^. Microneutralization Assay (MNA) was employed using Wild-Type SARS-COV-2 strain (Victoria isolate 01/2020). Pseudo virus Neutralization Assay was employed using Vascular Stomatis Virus expressing the SARS-COV-2 Spike protein and reporter gene Luciferase. The MNA was conducted at Translational Health Science and Technology Institute (THSTI), Faridabad, India and PNA was conducted at NEXELIS, Quebec, Canada; both are Coalition for Epidemic Preparedness Innovation (CEPI-network) labs. The nAb testing was conducted as per methods described previously^2^. Conversion factors have been established by both the laboratories to enable conversion of the Neutralization Titer (NT_50_) values to WHO-International Standard (NIBSC-20/136) and report the (NT_50_ values in International Units/mL^3^. MNA values were divided by 4.064 and PNA values were divided by 1.875 to obtain the titers in IU/mL when required for comparison. Geometric mean titers were calculated at scheduled time-points and fold Rise from the pre-vaccination values were calculated along with GMFR. Sera samples that did not demonstrate minimum 50% neutralization of the virus at the initial dilution i.e. the of the assay, titers were assigned as LLOQ/2. For key GMT/GMC values, 95% Confidence Intervals (95%CI) were also calculated. All the subject sera samples were tested for nAb titers using PNA method in Phase I/II and Phase II study. The subjects that had PNA titers below the LLOQ were considered as sero-negative for analysis while subjects with PNA titers above LLOQ were considered as sero-positive for analysis. Seroconversion rate for anti-RBD IgG concentration and nAb titers were calculated as follows: For sero-negative subjects, ≥4‑fold rise in IgG or nAb titers was considered to be seroconverted while for sero-positive subjects, ≥2‑fold rise in IgG or nAb titers was considered to be seroconverted.
4. Cellular immune responses were assessed in a subset of subjects in terms of cytokine secretion in TrueCulture^®^ tubes^4^. Whole blood samples from the subjects were incubated in growth medium in TrueCulture^®^ tubes coated with SARS-COV-2 Peptides (supplied by Myriad Inc, Texas, USA). The Peripheral Blood Mononuclear Cells present in the blood are stimulated by the peptides that resemble the vaccine antigen and in response to the stimulation the cells produce various cytokines. The culture supernatants were recovered and 2 cytokines; Interferon-gamma and Interlukin-4, secreted in the supernatant were measured by standard ELISA kits supplied by Becton Dickinson, USA. The whole blood stimulation was done at the clinical trial sites and the cytokine ELISA was conducted at Dang’s Lab. Cytokine values in pictogram/mL were reported for the sera samples and arithmetic averages were calculated for each cohort.
5. **Institutional Ethics Committees (IECs) of 25 study sites for phase-2/3 (BECT072) study**

| **S.No** | **Ethics Committee Name** | **Approval Status** | **EC Approval date** | **EC Registration No.** |
| --- | --- | --- | --- | --- |
| 1 | IEC-Mysore Medical College and Research Institute, Irwin Road, Mysuru 57000, Karnataka, India | Approved | 08-10-2021 | ECR/134/Inst/KA/2013/RR-19 |
| 2 | Institutional Ethics Committee JSS Medical College, JSS Medical College and Hospital, Sri Shivarathreeshwara Nagar, Mysuru - 570015, Karnataka, India | Approved | 05-10-2021 | ECR/387/Inst/KA/2013/RR-19 |
| 3 | Ethics Committee Guru Teg Bahadur Hospital Dilshad Garden, Delhi - 110095 India | Approved | 06-10-2021 | ECR/510/Inst/DL/2014/RR-20 |
| 4 | Institutional Ethics Committee, KLE University KLE University KLE Dr.PK Hospital and MRC, Nehru Nagar, Belagavi 590010 Karnataka, India. | Approved | 26-10-2021 | ECR/211/Inst/KA/2013/RR-19 |
| 5 | Shubham Sudbhawana Superspeciality Hospital - Ethics Committee, B 31/80, 23B - Bhogabeer, Lanka, Varanasi - 221005, Uttar Pradesh, India | Approved | 09-10-2021 | ECR/667/Inst/UP/2014/RR-20 |
| 6 | Institutional Ethics Committee, Government Medical College and Hospital, Government Medical College Medical Square, Hanuman Nagar Nagpur 440003, Maharashtra, India | Approved | 25-10-2021 | ECR/43/Inst/MH/2013/RR-19 |
| 7 | IEC NIMS University, Hotam administrative block, Delhi- Jaipur highway, Jaipur- 303121, Rajasthan, India. | Approved | 04-10-2021 | ECR/665/Inst/RJ/2014/RR-17 |
| 8 | Sant Dnyaneshwar Medical Education & Research Centre - Institutional Review Board, 695/A, Sadashiv Peth, Opp. Vijay Talkies, Laxmi Road, Pune 411030, Maharashtra | Approved | 27-10-2021 | ECR/127/Inst/MH/2013/RR-19 |
| 9 | ETHICS COMMITTEE, St. Theresa's Hospital, Sanath Nagar, Opp. Erragada Raitu Bazar, Hyderabad 500018, Telangana, India | Approved | 01-10-2021 | ECR/230/Inst/AP/2013/RR-19 |
| 10 | IEC, Mahatma Gandi Institute Of Medical Sciences, Sewagram, Wardha 442102, Maharshtra, india | Approved | 23-10-2021 | ECR/47/Inst/MH/2013/RR-19 |
| 11 | Ethics Committee Dy.Patil Vidyapeeth, Pimpri,  Pune - 411018, Maharashtra, India | Approved | 07-12-2021 | ECR/361/Inst/MH/2013/RR-19 |
| 12 | IRB Christian Medical college, Office of Research, 1st floor, Carman Block, Bagayam, Vellore 632002, Tamilnadu, India. | Approved | 30-10-2021 | ECR/RENEW/INST/2019/3526 |
| 13 | Institutional Ethics Committee for ESIC Medical College And Hospital NH-3, NIT, Behind BK Hospital, Faridabad 121001, Haryana, India | Approved | 16-10-2021 | ECR/1539/Inst/HR/2021 |
| 14 | IEC Prakhar Hospital Pvt Ltd, 8/219, Arya Nagar, Kanpur 208002, Uttar Pradesh, India. | Approved | 20-10-2021 | ECR/1017/Inst/UP/2017/RR-21 |
| 15 | Aakash Healthcare Institutional Ethics Committee,  Road No. 201, Sector 3, Dwarka, New Delhi - 110075, India | Approved | 09-11-2021 | ECR/1265/Inst/DL/2019 |
| 16 | Gullurkar Hospital Ethics Committee, Plot no. 20, Reshimbag Umred Road, Nagpur- 440009, Maharashtra, India | Approved | 18-11-2021 | ECR/1374/Inst/MH/2020 |
| 17 | Institutional Ethics Committee- Induss Hospital, 13-23-93/1, Krishnaveninagar, Gaddiannaram, Near Muncipal Office, Saroornagar, Ranga Reddy Dist., Hyderabad 500035, Telangana, India | Approved | 06-11-2021 | ECR/1606/Inst/TG/2021 |
| 18 | Institutional Ethics Committee- Jeevan Rekha Hospital, Dr. B.R. Ambedkar Road,Opp Civil Hospital, Belagavi- 590002, Karnataka, India | Approved | 06-11-2021 | ECR/1242/Inst/KA/2019 |
| 19 | Institutional Ethics Committee - Jawahar Lal Nehru Medical College, Kala Bagh Ajmer - 305001, Rajasthan, India | Approved | 22-11-2021 | ECR/1156/Inst/RJ/2018/RR-22 |
| 20 | Lifepoint Research- Ethics Committee, 145/1, Mumbai-Bangalore highway, Near Hotel Sayaji, Wakad, Pune - 411057, Maharashtra, India | Approved | 09-11-2021 | ECR/751/Inst/MH/2015/RR-21 |
| 21 | Penta-Med Ethics Committee,   Medipoint Hospitals Pvt. Ltd, 241/1, New D.P.Road, Near Sai Heritage,Pune - 411007, Maharashtra, India | Approved | 12-11-2021 | ECR/357/Inst/MH/2013/RR-20 |
| 22 | Unique Children's Hospital Ethics Committee, Hira Moti Fortune Opp. Police station, Pune Mumbai Road, Chinchwad, Pune-411019, Maharashtra, India | Approved | 16-12-2021 | ECR/1203/Inst/MH/2019 |
| 23 | CARE-IHEC for Faculty Research Chettinad Academy Of Research And Education, Rajiv Gandhi Salai (OMR),Kelambakkam, Kanchipuram - 603103, Tamil Nadu, India | Approved | 09-12-2021 | ECR/1589/Inst/TN/2021 |

**References**.

1.	NIBSC. Data Sheet -Research reagent for anti-SARS-CoV-2 Ab NIBSC code 20/130.

2.	Bewley KR, Coombes NS, Gagnon L, et al. Quantification of SARS-CoV-2 neutralizing antibody by wild-type plaque reduction neutralization, microneutralization and pseudotyped virus neutralization assays. *Nat Protoc* 2021; **16**(6): 3114-40.

3.	Mattiuzzo G, Bentley EM, Hassall M, et al. Establishment of the WHO International Standard and Reference Panel for anti-SARS-CoV-2 antibody. *World Health Organization* 2020; **60**.

4.	Petrovsky N, Harrison LC. Cytokine-based human whole blood assay for the detection of antigen-reactive T cells. *J Immunol Methods* 1995; **186**(1): 37-46.
